# Programmatic Effectiveness of a Pediatric Typhoid Conjugate Vaccine Campaign in Navi Mumbai, India

**DOI:** 10.1101/2022.10.26.22281529

**Authors:** Seth A. Hoffman, Christopher LeBoa, Kashmira Date, Pradeep Haldar, Pauline Harvey, Rahul Shimpi, Qian An, Chenhua Zhang, Niniya Jayaprasad, Lily Horng, Kirsten Fagerli, Priyanka Borhade, Debjit Chakraborty, Sunil Bahl, Arun Katkar, Abhishek Kunwar, Vijay Yewale, Jason R. Andrews, Pankaj Bhatnagar, Shanta Dutta, Stephen P. Luby

## Abstract

**Background:** The WHO recommends vaccines for prevention and control of typhoid fever, especially where antimicrobial-resistant typhoid circulates. In 2018 the Navi Mumbai Municipal Corporation (NMMC), implemented a TCV campaign. The campaign targeted all children aged 9-months through 14-years within NMMC boundaries (∼320,000 children) over 2 vaccination phases. The phase 1 campaign occurred from July 14-August 25, 2018 (71% coverage, ∼113,420 children). We evaluated the campaign’s programmatic effectiveness in reducing typhoid cases at the community level.

**Methods:** We established prospective, blood culture-based surveillance at 6 hospitals in Navi Mumbai, offering blood cultures to children presenting with fever for at least 3 days. We employed a cluster-randomized test-negative design to estimate the effectiveness of the vaccination campaign on pediatric typhoid cases. We matched culture-confirmed typhoid cases with up to 3 culture-negative controls by age and date of blood culture and assessed community vaccine campaign phase as an exposure using conditional logistic regression.

**Results:** Between September 1, 2018–March 31, 2021, we identified 81 typhoid cases and matched these with 238 controls. Cases were 0.44 times as likely to live in vaccine campaign communities (campaign effectiveness, 56%, 95%CI: 25%-74%, p=0.002). Cases ≥ 5-years-old were 0.37 times as likely (95% CI: 0.19-0.70; p-value = 0.002) and cases during the first year of surveillance were 0.30 times as likely (95% CI: 0.14-0.64; p-value = 0.002) to live in vaccine campaign communities.

**Conclusions:** Our findings support the use of TCV mass vaccination campaigns as effective population-based tools to combat typhoid fever.

**Summary:** In 2018, the Navi Mumbai Municipal Corporation conducted a typhoid conjugate vaccine campaign in half of its communities. Utilizing a test-negative design, we estimate that this campaign reduced typhoid risk by 56% (25-74%) in vaccinated communities compared to non-campaign communities.

## INTRODUCTION

Typhoid fever, caused by *Salmonella enterica* subspecies *enterica* serovar Typhi (*S*. Typhi), remains the cause of a significant burden of disease, with an estimated over 11 million cases and 116,000 deaths in 2017, the majority of which occurred in children under 15 years of age [1]. In the mid-20^th^ century typhoid fever mortality dropped significantly, from upwards of 12% in the pre-antibiotic era [2], to approximately one percent in most areas after the introduction of effective antibiotics [3, 4]. Unfortunately, antimicrobial-resistant *S*. Typhi is on the rise, especially in Asia [5, 6], and is associated with increased severity of disease [7–9].

In conjunction with water, sanitation, and hygiene (WASH) interventions, the World Health Organization (WHO) recommends vaccines as an important tool in typhoid prevention and control [10]. The WHO prequalified the first typhoid conjugate vaccine (TCV), Typbar-TCV® (Bharat Biotech International Limited, India), which comprises the Vi polysaccharide conjugated to tetanus toxoid, in December 2017 for use in children as young as 6 months old [10]. A second TCV (TYPHIBEV®, Biological E. Limited, India) was prequalified in December 2020 [11]. Earlier typhoid vaccines, Ty21a and the unconjugated Vi polysaccharide (ViPS) vaccine, are not recommend for infants and the ViPS vaccine is not as immunogenic as TCVs [10]. In multiple studies, including randomized controlled trials, ranging from infants to adults, TCVs have shown 79-95% efficacy in preventing typhoid fever [12–18], and earlier analysis from this project measured a vaccine effectiveness of 82% in our sampled population [19]. Additionally, TCV can safely be co-administered with other routine vaccines without immune interference or increased adverse events [20], making it ideal for inclusion in a country’s routine immunization schedule.

Navi Mumbai, India is an urban municipality outside of Mumbai administered by the Navi Mumbai Municipal Corporation (NMMC). Prior observations suggested a high burden of typhoid in children [21]. In 2018, the NMMC implemented the first phase of a public-sector pediatric TCV campaign [22]. The campaign targeted all children aged 9-months through 14-years living within NMMC boundaries (∼320,000 children) over the course of 2 vaccination phases (each ∼160,000 children). The selection of areas to include in the two phases of the campaign were initially developed as a single step wedge design randomized control trial in collaboration with NMMC. The first phase of the TCV mass vaccination campaign occurred over the course of 6 weeks between 14 July and 25 August 2018, and reached an estimated 113,420 children (71% of the target population); each child received one dose of Typbar-TCV®. At the time of vaccination, each family received a TCV vaccination card to keep as a part of their medical record. The second, delayed vaccination campaign was planned for 2020, but was postponed due to the COVID-19 pandemic and has not yet been conducted. The first confirmed case of COVID-19 was reported in India on January 30, 2020 [23] and a subsequent countrywide lockdown started on March 25, 2020.

While previous randomized clinical trial data provide safety and efficacy data, only in Pakistan has TCV effectiveness been evaluated in a public sector, outbreak setting [15]. Our study provides the first evaluation of a public sector mass vaccination campaign using TCV in a non-outbreak setting. This information may be useful to key stakeholders and policymakers as decisions are made on whether to introduce TCV into routine immunization programs through immunization campaigns in *S*. Typhi-endemic countries.

## METHODS

### Setting

NMMC’s administrative boundaries are divided into 22 areas of approximately even population size, demarcated by the catchment areas of the city’s 22 Urban Health Posts (UHPs). In conjunction with NMMC, we stratified areas based on proportion of the population living in slums (stratum 1, <10%; stratum 2, 10%-70%; and stratum 3, >70%) [22], and randomly assigned areas either to vaccine campaign communities, which underwent the mass vaccination campaign in 2018, or delayed vaccine campaign communities, which were to be included in the second phase of the vaccination campaign (Figure 1). Children aged 9-months through 14-years were eligible for the vaccination campaign, amounting to an estimated 320,000 children within the NMMC administrative boundaries, and an estimated 160,000 children targeted for immunization in the first phase of the campaign.

**Figure 1.**
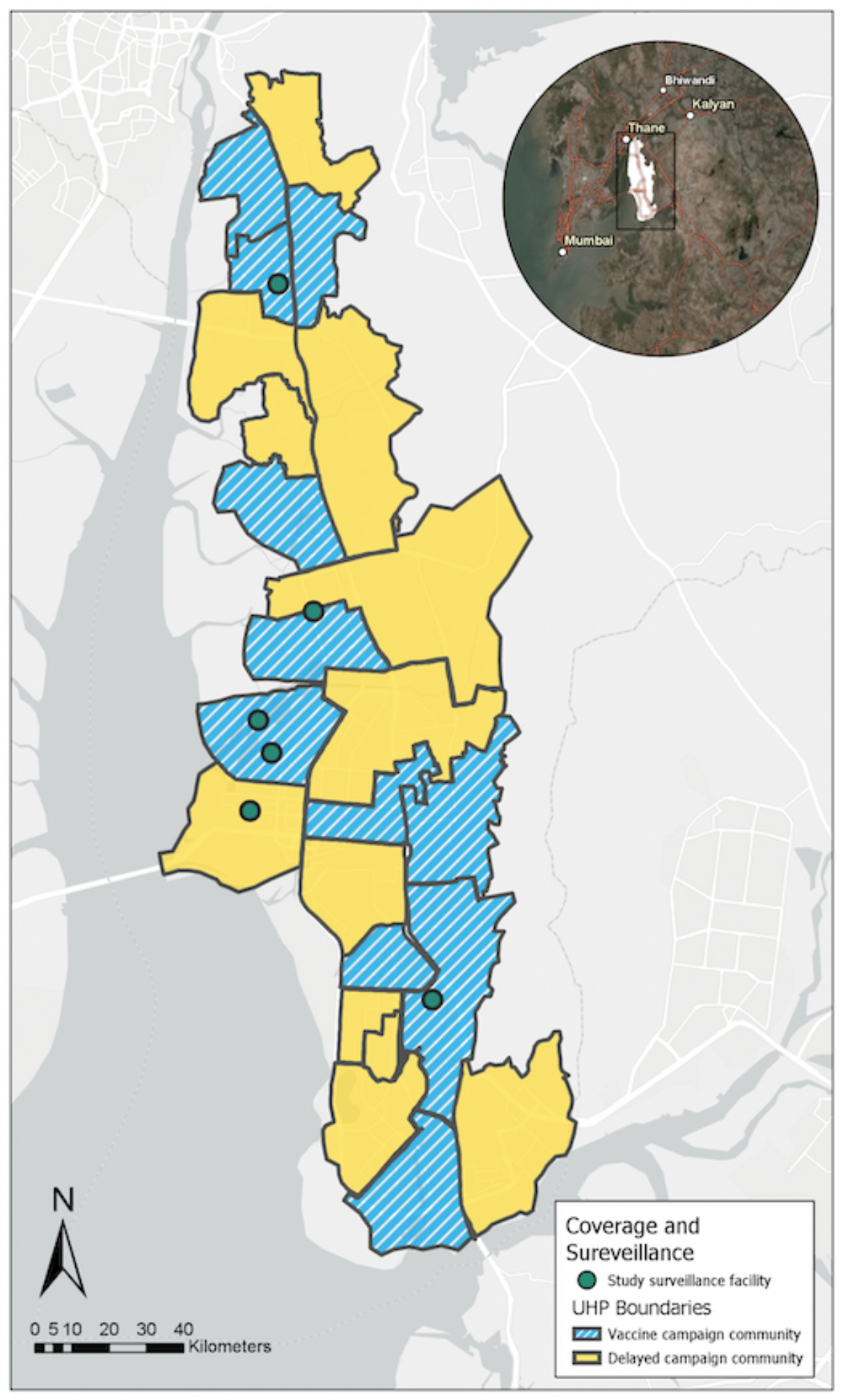
Map of vaccine campaign and delayed campaign communities. Map of vaccine campaign communities (blue stripes) and delayed campaign communities (yellow solid) as demarcated by the borders of Navi Mumbai Municipal Corporation’s 22 Urban Health Posts. Hospitals that participated in healthcare facility blood culture surveillance marked by green circles.

### Design

#### Original design and statistical analysis plan

We planned to estimate the incidence of blood culture-confirmed typhoid among children eligible to receive the vaccine in the vaccine campaign communities and compare this to the delayed campaign communities (https://clinicaltrials.gov/ct2/show/NCT03554213), following a single step wedge design. We intended to deploy hybrid surveillance that adjusted the crude typhoid fever incidence for healthcare utilization [24], utilizing prospective, blood culture-based surveillance at 6 hospitals and a continuous community survey. The COVID-19 pandemic radically altered healthcare seeking behavior in Navi Mumbai and necessitated the suspension of the continuous community survey to minimize risk to fieldworkers. But the healthcare facility blood culture surveillance continued (September 1, 2018 through March 31, 2021) providing community-level blood culture data.

#### Revised design and statistical analysis plan

To evaluate the effectiveness of the mass vaccination campaign in the setting of radically altered and reduced healthcare seeking, we implemented a test-negative case-control design [25]. Among pediatric patients presenting with a febrile illness to affiliated healthcare sites, we identified cases as being blood culture positive for *S*. Typhi and controls as being blood culture negative for *S*. Typhi, and we then examined home address in a vaccine campaign community or a delayed campaign community as the exposure of interest.

#### Inclusion Criteria

We included children within the age range of eligibility for the vaccination campaign (9-months through 14-years), and through 16-years to account for aging during the follow up period, who presented or were sent to a hospital sentinel site with a febrile illness (>3 days of fever within the past 7 days without upper respiratory tract symptoms and/or vesicular rash). The hospital sites were D.Y. Patil Medical College and Hospital, Nerul; Dr. Yewale Multispecialty Hospital for Children, Vashi; Mahatma Gandhi Memorial (MGM) New Bombay Hospital, Vashi; Mathadi Hospital Trust, Koparkharaine; NMMC General Hospital, First Referral Unit (FRU), Vashi; and Rajmata Jijau Hospital, NMMC, Airoli. We offered free blood culture to all participants who met the inclusion criteria. Participants were included for 33 months (September 1, 2018 – March 31, 2021).

#### Exclusion Criteria

Participants were excluded if no result was recorded for their blood culture or if they did not have a place of residence listed or listed a location outside of NMMC boundaries.

### Data Analysis

We matched each case with up to 3 controls on age (+/−12 months) and time of enrollment (+/−28 days) to account for age-related differences in typhoid incidence [1, 26, 27] and the seasonality of typhoid fever in the region [28]. We attempted to match on shorter time intervals of enrollment, but these failed to match all cases (R MatchIt Package,R version 4.0.4).

We performed conditional logistic regression (clogit function from R “survival” package, R version 4.0.4) to estimate the odds that a blood culture-positive *S*. Typhi case resided in the vaccine campaign communities versus the delayed vaccine campaign communities. We adjusted the model for participant sex and education group (whether respondent has more than primary school education or not). Programmatic campaign effectiveness was calculated as (1 – Odds Ratio) x 100.

We performed stratified analyses by participant age group (<5 years of age versus ≥5 years of age), to assess age-specific effectiveness of the vaccine, and by time since vaccine campaign (≤ 365 days [September 1, 2018 – August 31, 2019] versus > 365 days [September 1, 2019 – March 31, 2021] since phase 1 of the campaign) to evaluate any differences in effectiveness between the first and second-third years after the campaign. These stratified analyses were both tested for the significance of the difference by interaction, as calculated by an ANOVA test for the null versus an interaction model, where the interaction terms were “(study community)*(age strata)” in the age stratified analysis and (study community)*(time since vaccine campaign strata)” in the time since vaccine campaign stratified analysis, where study community is defined by vaccine campaign communities versus delayed vaccine campaign communities.

### Ethics Statement

We obtained written informed consent from adult caregivers and verbal assent from pediatric participants aged >12 years. The study protocol was approved by MGM New Bombay Hospital IRB, Vashi, India; Institutional Ethics Committee, Indian Council of Medical Research – National Institute of Cholera and Enteric Diseases (No. A-1/2020-IEC); WHO Research Ethics Review Committee (ERC.0002923); and Stanford University IRB (IRB-39627).

## RESULTS

During September 1, 2018 – March 31, 2021 (33 months), we enrolled 2,953 participants who met the fever definition (Figure 2). There were a total of 81 blood-culture positive cases of typhoid. After matching on date of test (+/−28 days) and age (+/−12 months), we included 319 participants in our analysis (Table 1, Figure 1). Among the 81 blood culture-positive cases of *S*.Typhi, 36 were from the vaccine campaign communities and 45 were from the delayed vaccine campaign communities. There were 238 blood-culture negative controls, 153 from the vaccine campaign communities and 85 from the delayed vaccine campaign communities. The mean age of the cases was 6.9 years and the matched controls 6.8 years old. Females represented 40% of cases and 47% of matched controls.

**Figure 2.**
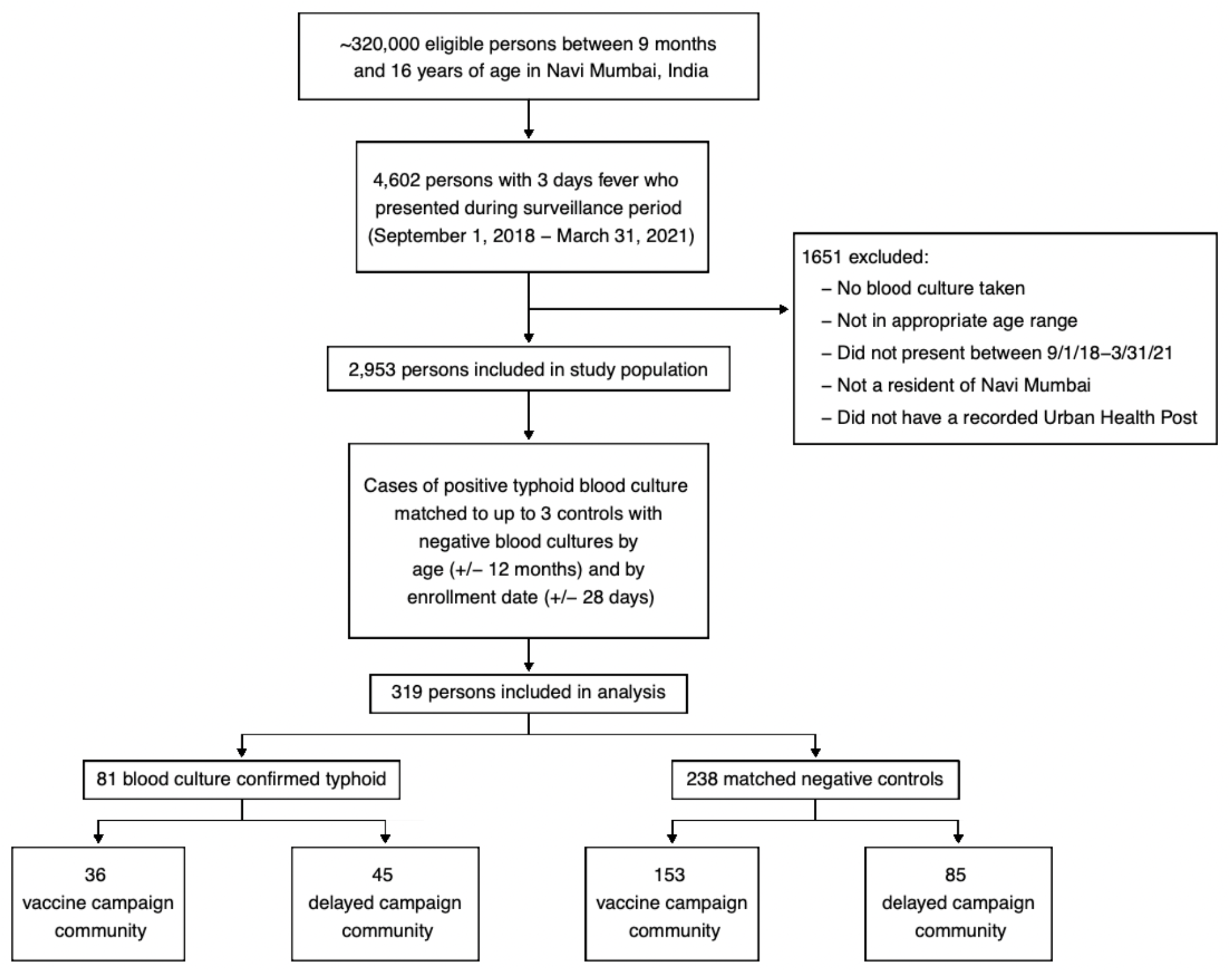
Trial Profile. Flow diagram depicting total estimated eligible participant population in Navi Mumbai (∼320,000), number of participants reported by healthcare facility blood culture surveillance (4,602), number of participants with complete records (2,953), number of participants included in analysis after matching (319), and blood culture positive and negatives included in analysis.

**Table 1:** Demographic characteristics of child participants and adult respondents.

|  | Blood-culture<br>positive cases<br>(N=81) | Blood-culture negative<br>controls<br>(N=238) | P-value |
| --- | --- | --- | --- |
| Child Age |  |  |  |
| Mean (SD) | 6.85 (3.54) | 6.77 (3.54) | 0.9 |
| Median [Min, Max] | 7.00 [1.00, 15.0] | 7.00 [1.00, 15.0] |  |
| Child Gender |  |  |  |
| Male | 49 (60.5%) | 127 (53.4%) | 0.3 |
| Female | 32 (39.5%) | 111 (46.6%) |  |
| Adult Respondent Gender |  |  |  |
| Male | 38 (46.9%) | 122 (51.3%) | 0.5 |
| Female | 42 (51.9%) | 113 (47.5%) |  |
| Missing | 1 (1.2%) | 3 (1.3%) |  |
| Adult Respondent Age |  |  |  |
| Mean (SD) | 34.9 (7.43) | 34.6 (8.66) | 0.7 |
| Median [Min, Max] | 35.0 [19.0, 59.0] | 34.0 [18.0, 85.0] |  |
| Missing | 0 (0%) | 1 (0.4%) |  |
| Adult Respondent Education Level |  |  |  |
| Primary school certificate (through Grade 4) or less | 6 (7.4%) | 32 (13.4%) | 0.1 |
| Middle school certificate (Grade 5 - 10) | 35 (43.2%) | 112 (47.1%) |  |
| High school certificate (Grade 11 - 12) | 14 (17.3%) | 45 (18.9%) |  |
| Bachelors degree or greater | 25 (30.9%) | 49 (20.6%) |  |
| Don't know or refused | 1 (1.2%) | 0 (0%) |  |

The unadjusted odds ratio (OR) that culture-confirmed typhoid cases resided in the vaccine campaign communities versus in the delayed vaccine campaign communities was 0.44 (95% CI: 0.27-0.74; p-value = 0.002). The adjusted odds ratio (aOR) was 0.44 (95% CI: 0.26-0.75; p-value = 0.003), indicating that the effectiveness of the mass vaccination campaign was 56% (95% CI: 25-74%) in the vaccine campaign communities (Table 2).

**Table 2.**
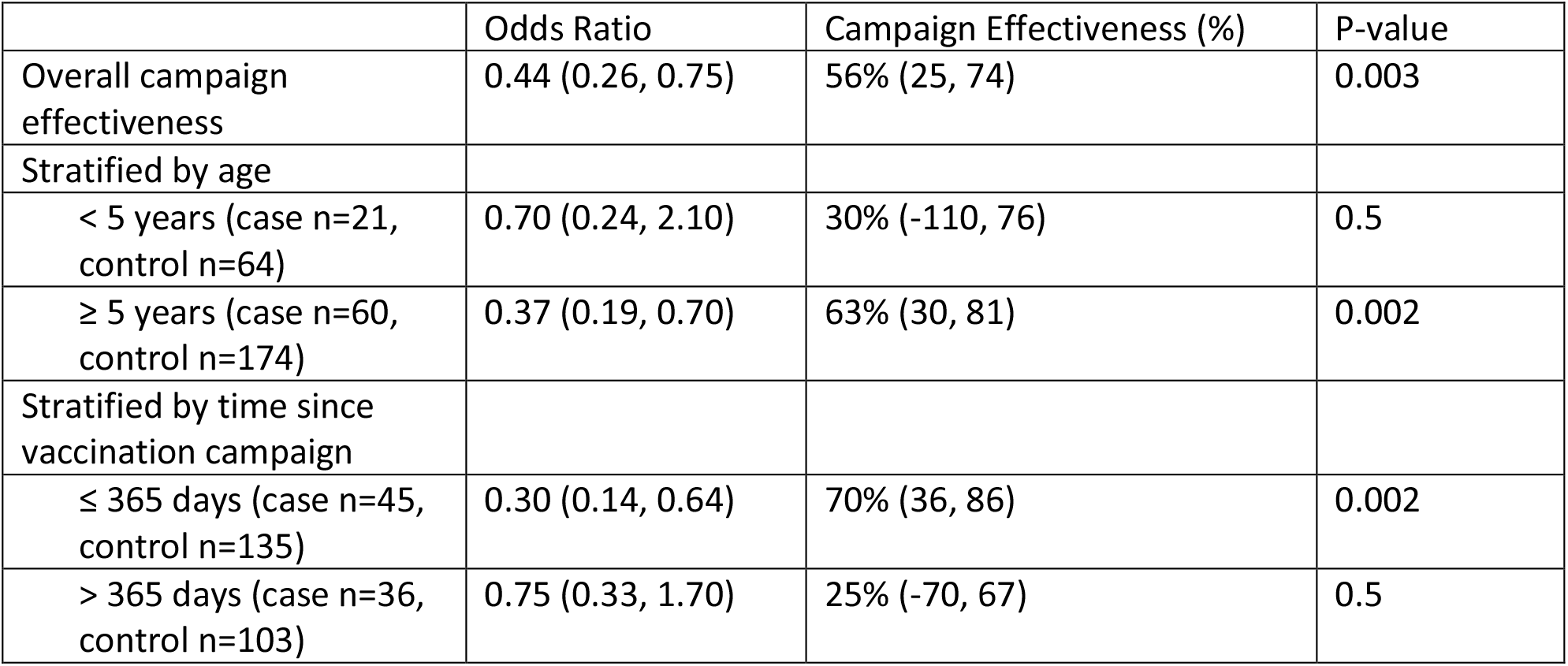
Overall and Stratified Campaign Effectiveness.

|  | Odds Ratio | Campaign Effectiveness (%) | P-value |
| --- | --- | --- | --- |
| Overall campaign effectiveness | 0.44 (0.26, 0.75) | 56% (25, 74) | 0.003 |
| Stratified by age |  |  |  |
| < 5 years (case n=21, control n=64) | 0.70 (0.24, 2.10) | 30% (-110, 76) | 0.5 |
| ≥ 5 years (case n=60, control n=174) | 0.37 (0.19, 0.70) | 63% (30, 81) | 0.002 |
| Stratified by time since vaccination campaign |  |  |  |
| ≤ 365 days (case n=45, control n=135) | 0.30 (0.14, 0.64) | 70% (36, 86) | 0.002 |
| > 365 days (case n=36, control n=103) | 0.75 (0.33, 1.70) | 25% (-70, 67) | 0.5 |

When stratified by participant age group (< 5 years of age versus ≥ 5 years of age), amongst the under 5 years participants, cases were 0.70 times as likely to live in vaccine campaign communities compared with delayed vaccine campaign communities (campaign effectiveness, 30%, 95% CI: -110-76; p-value = 0.5), while cases aged 5 years and older were 0.37 times as likely to live in vaccine campaign communities (campaign effectiveness, 63%, 95% CI: 30-81; p-value = 0.002). When stratified by time since campaign (≤ 365 days versus > 365 days since campaign), cases occurring within the first year after the campaign were 0.30 times as likely to live in vaccine campaign communities (campaign effectiveness, 70%, 95% CI: 36-86; p-value = 0.002), while cases identified during the second-third year after the campaign were 0.75 times as likely to live in vaccine campaign communities (campaign effectiveness, 25%, 95% CI: -70-67; p-value = 0.5).

## DISCUSSION

The TCV campaign successfully vaccinated an estimated 113,420 children in 6 weeks. Typhoid fever cases were 56% less likely to reside in vaccine campaign communities than in delayed vaccine campaign communities. When stratified by age, older case-patients (>5 years old) were 63% less likely to reside in vaccine campaign communities than delayed vaccine campaign communities, but this difference was not significant when tested for the significance of the difference by interaction. Additionally, when stratified by time since vaccine campaign surveillance, typhoid fever cases were 70% less likely to live in the vaccine campaign communities during the first year of surveillance, but this difference was also not significant when tested for the significance of the difference by interaction.

The overall programmatic effectiveness of the vaccination campaign on the reduction of typhoid in the intervention community was consistent with 71% estimated vaccine coverage of an 82% effective vaccine. Our crude and adjusted overall campaign effect odds ratios were similar, indicating that adjustment for participant sex and respondent education had little impact, and these variables were unlikely to be strong confounders.

To minimize risk to project fieldworkers from the COVID-19 pandemic, we halted the continuous community survey. Since the original study design and statistical analysis plan did not allow for this significant impact on data collection, and given the effect of the marked change in healthcare utilization due to the COVID-19 pandemic, we could not follow the prespecified study design. Instead, utilizing the available blood culture data, we employed a test-negative design to analyze our results [29, 30]. This allowed us to evaluate the overall impact of the vaccine campaign on the reduction of typhoid cases between the intervention communities versus the delayed vaccine campaign communities. This study design, modeled off the popular test-negative design used in influenza [31], rotavirus [32], and COVID-19 [33, 34] vaccine effectiveness studies, has been shown to be concordant with vaccine effectiveness results from randomized controlled trials [25, 32]. The cluster randomized test-negative design has also been utilized in the evaluation of dengue control interventions [35]. Recently, the WHO issued guidance recommending the use of the test-negative design for COVID-19 vaccine effectiveness studies due to logistical ease and reduction of health utilization bias [33, 34].

In addition to the unpredictable impact of the COVID-19 pandemic, this study is limited by utilizing *S*. Typhi blood culture as the outcome measure for test-positive cases versus test-negative controls. Blood culture sensitivity in detecting *S*. Typhi infection is estimated to be between 55-65% [1, 36] and is dependent on the blood volume collected [36], for which we do not have data. As such, blood culture positivity as an outcome measure may result in underreporting of test-positive cases and loss of statistical power, but this would be expected to affect both vaccine campaign and delayed vaccine campaign communities and so would not bias the estimate of campaign effectiveness. For now, blood culture remains the standard outcome utilized by evaluations of interventions to combat *S*. Typhi infection in controlled trials and public health programs, and *S*. Typhi surveillance [1, 5, 6, 9, 13–15, 17, 18].

There are a number of descriptions in the literature of programmatic or outbreak implementation of typhoid vaccines [15, 37–39]. However, formal assessment is limited and many lack a systematically evaluated population control group, limiting generalizability [15, 37–39]. Our results represent a metropolitan introduction of typhoid vaccine in which we systematically evaluated the overall campaign effectiveness with control communities that had not yet undergone a TCV immunization campaign. As compared to previous generation typhoid vaccines, unconjugated Vi polysaccharide and Ty21a, TCVs induce consistently higher levels of protective efficacy and are indicated for use in younger children [10]. TCVs provide more durable immune responses than do unconjugated Vi polysaccharide vaccine, and are expected to have longer durations of protection than prior generation typhoid vaccines [10, 16]. This study provides further evidence that TCVs are effective when rolled out in the form of a municipal program.

Although the campaign did reduce typhoid, some children who lived in the intervention community still developed typhoid, even some of those children who were vaccinated [19]. Effective communication to stakeholders and policymakers should include the caveat that while programmatic implementation of TCV in children and adolescents will reduce typhoid incidence, it is not likely to interrupt all transmission and eliminate the disease for several reasons [14]. First, although highly effective, typhoid vaccines are not 100% effective, as has been reported from individually randomized trials [12–18]. Second, adults who are susceptible to typhoid, are not usually included in typhoid immunization campaigns or programs. These adults sometimes develop typhoid fever and transmit *S*. Typhi. Even the targeted age groups are unlikely to be fully reached by vaccine campaigns – in this study vaccine coverage was an estimated 71%. Finally, vaccination will not cure *S*. Typhi chronic carriers, who will remain a reservoir for future transmission. Thus, public health planners should be prepared to communicate that TCV reduces but does not eliminate risk, and increased attention to providing food and drinking water not contaminated with feces will be important to reduce typhoid-associated morbidity and mortality.

Our findings support the use of TCV mass vaccination campaigns as an effective population-based tool to reduce the pediatric public health burden of typhoid fever, as part of the introduction of TCV into routine immunization programs. This evaluation adds to the body of research supporting TCV campaigns by demonstrating these campaigns can be effective at the population level, while prior modeling suggests they may be highly cost-effective [40]. Additionally, our experience is in the setting of atypical population healthcare utilization or low reported typhoid incidence, the test-negative design is useful in estimating programmatic campaign effectiveness. Our data adds to the growing evidence that stakeholders can use to support decisions on TCV introduction to reduce the burden of typhoid fever.

## Data Availability

All data produced in the present study are available upon reasonable request to the authors

## ACKNOWLEDGEMENTS

We thank the following organizations and individuals for their contributions to this evaluation: the Navi Mumbai Municipal Corporation leadership and staff, the Government of India Ministry of Health and Family Welfare Universal Immunization Program, State of Maharashtra Department of Public Health and Family Welfare, Indian Academy of Pediatrics Navi Mumbai Chapter, Bharat Biotech International Limited, the Indian Council of Medical Research, WHO-India National Public Health Surveillance Project, the Navi Mumbai Municipal Corporation Hospital (Dr. Savita Daruwalla), D.Y. Patil Medical College and Hospital (Dr. Rajesh Rai, Dr. Keertana Shetty), Mathadi Trust Hospital (Dr. Divya Warrier), MGM New Bombay Hospital Vashi (Dr. Jeetendra Gavhane, Dr. Shalini Yadav), Dr. Yewale Multi-Specialty Hospital for Children (Dr. Dhanya Dharmapalan), Grant Government Medical College (Dr. Nilma Hirani), Dr. Joshi’s Central Clinical Microbiology Laboratory, Vashi (Dr. Shrikrishna Joshi), Rajmata Jijau Hospital, NMMC, Airoli (Dr. Varsha Rathod), and the Centers for Disease Control and Prevention, Atlanta, Georgia (Dr. Kathleen Wannemuehler, Benjamin Nygren, and Matt Mikoleit).

## DISCLAIMER

The findings and conclusions in this report are those of the authors and do not necessarily represent the official position, policies, or views of the United States Centers for Disease Control and Prevention, or the World Health Organization.

